# Combining phenotypic similarity and network propagation to improve performance and clinical consistency of rare disease diagnosis

**DOI:** 10.64898/2026.02.15.26346357

**Authors:** Maroua Chahdil, Carolina Fabrizzi, Marc Hanauer, Caterina Lucano, Ana Rath, David Lagorce, Laurent Tichit

## Abstract

Achieving timely diagnosis for rare diseases remains challenging due to, among others, phenotypic heterogeneity and incomplete clinical data. While the Solve-RD project developed a phenotype-based gene prioritisation method, this approach did not account for the clinical consistency among related diseases in Orphanet’s hierarchical classifications.

We present a phenotype-based computational pipeline that ranks candidate ORPHAcodes based on patient phenotypes. The pipeline computes patient-disease similarity using asymmetric semantic aggregation of Human Phenotype Ontology terms, filtering subsumed terms and incorporating Orphanet frequency annotations. Evaluated on 139 expert curated Solve-RD cases representing 78 distinct ORPHAcodes, our methodology outperformed the established Solve-RD baseline method, achieving a harmonic mean rank of 4.64 for confirmed diagnoses (versus 7.97) and retrieving the correct suspected rare disease within the top 10 positions for 39% of patients (versus 29%). We then explore a disease similarity network using Random Walk with Restart to generate ranked candidate lists. Two complementary experiments demonstrate that RWR-ranked candidates exhibited improved clinical consistency, reflected by their proximity within the Orphanet nomenclature of rare diseases. This approach provides more interpretable and actionable differential diagnosis hypotheses to guide clinical decision-making

**Author summary:** Many patients with rare diseases face prolonged diagnostic delays due to the extreme heterogeneity of rare disorders associated with the variability of their clinical manifestations, which complicates interpretation and requires structured phenotypic representations and expert knowledge. We developed a computational pipeline that compares patients’ phenotypes with those documented for rare diseases in the Orphanet database. Rather than relying solely on direct matching of clinical signs and symptoms, our approach leverages relationships between diseases by propagating information through a network connecting patients and diseases. Testing on 139 cases from the European Solve-RD project, our method improved identification of correct diagnoses and generated more clinically coherent candidate lists by accounting the Orphanet nomenclature. This work provides a methodology dedicated to assisting clinicians in developing diagnostic hypotheses for rare diseases.

## Introduction

A disease is defined in Europe as a Rare Disease (RD) when the number of affected patients equal or fewer than 1 in 2,000 subjects [1]. According to Orphanet (www.orpha.net), more than 6,000 RDs have been identified worldwide, 72.2% of which have a genetic origin. However, despite this predominance of genetic etiologies, around 35% of RDs still lack an identified causative gene [2]. Establishing an accurate clinical diagnosis of RD remains particularly challenging because these diseases often affect multiple organ systems or physiological functions [3]. Consequently, patients often experience prolonged diagnostic delays and may receive initial misdiagnoses, with some remaining undiagnosed. Several large-scale initiatives aim to improve diagnostic outcomes for RD, including the UK 100,000 Genomes Project [4] and the European Solve-RD project (https://solve-rd.eu/) [5].

Within the Solve-RD project, a phenotype-based semantic similarity pipeline, employing the Human Phenotype Ontology (HPO) [6], was developed to guide variant prioritization and generate diagnostic hypotheses [7], alongside efforts to uncover molecular causes in patients remaining undiagnosed after whole exome sequencing [8,9]. The computational package *RunSolveRD* (RSD) [10] was developed to compute similarity between clinical entities based on phenotypic annotations. Nevertheless, RSD exhibits sensitivity to sparsely annotated diseases, resulting in variability in candidate gene lists, and its closed-source framework limits extensibility, maintenance and adaptation. Over the last decade, various graph-based methods and tools have been introduced to exploit ontology-based semantic similarity. These methods fall into three categories: (i) graph-based methods that compute ontology-based semantic similarity, such as the Wang method [11] ; (ii) likelihood-ratio approaches, like RDmaster [12] (8) and PheLR [13], which combine HPO annotations with epidemiological data to compute phenotypic evidence for candidate diseases; and (iii) network-based methodologies such as RDmap [14] that model relationships between diseases and patients. Network propagation methods like Random Walk with Restart (RWR) can handle sparse or noisy phenotype annotations by diffusing information across neighbouring disease nodes [15]. More recent tools such as Phen2Disease [16] and PhenoDP [17] further integrate bidirectional matching or learned embeddings to prioritise candidate diseases or genes.

Despite these methodological advances, several aspects remain underexplored in phenotype-driven RD diagnostics, notably the integration of Orphanet’s hierarchical rare disease classifications structure. In this study, we present a decision-support approach that combines group-wise semantic similarity measures with network propagation using RWR. Our method computes phenotypic similarity between patients’ clinical signs and those annotated in each ORPHAcode (Orphanet’s unique and time-stable RD identifiers), while incorporating estimated phenotype frequency and disease classification level in order to improve both ranking performance and clinical coherence of diagnostic hypotheses.

## Materials and Methods

### Orphanet nomenclature and clinical consistency Data used

#### The Human Phenotype Ontology (HPO)

HPO [6] provides a standardised vocabulary of phenotypic abnormalities encountered in human disease. For this study, we used the HPO 2025-03-03 release which embedded 14570 unique HPO terms.

#### Orphanet data

We employed a reference collection based on the ’*Phenotypes Associated with Rare Disorders*’ May 2025 release from Orphadata.com, which includes 4,319 disorder-level ORPHAcodes, manually curated and annotated with HPO terms. At Orphanet, the Phenotypes Database Manager is responsible to systematically review the literature to identify disease-associated phenotypic abnormalities, map them to the most appropriate HPO terms, and classify each sign by frequency of occurrence in the disease population. Indeed, for each HPO term associated with a rare disease Orphanet assigns six frequency categories estimated from qualitative prevalence in affected populations: obligate (100%), very frequent (99-80%), frequent (79-30%), occasional (29-5%), very rare (4-1%) and excluded (0%). When no suitable HPO term exists, the closest term is used and a request for a new term is submitted to the HPO team. Herein this dataset for our study, to ensure a consistent unit of representation across disease annotations and patient diagnosis aligned to the Orphanet nomenclature principles, all implicated ORPHAcodes were harmonised to their reference ORPHAcode Disorder level.

#### Solve-RD case cohort subset

To evaluate and validate the proposed pipeline, we used the anonymised “solved patients” data from the Solve-RD project, stored in the Phenopackets format, a computer-readable standard compliant with GA4GH specifications (https://www.ga4gh.org/). Between 2018 to 2023, clinicians affiliated with the Solve-RD consortium annotated medical records of patients suffering from undiagnosed RDs, using a minimum of five HPO clinical signs. Diagnostics information and HPO annotations were extracted directly from the corresponding phenopacket files.

Within the Solve-RD dataset, a subset of 817 patients had a confirmed diagnosis mapped to a known ORPHAcode with some could be associated with multiple patients. An Orphanet medical expert manually reviewed these cases, and selected 139 patients whose phenotypic profiles were consistent with the expected phenotype of the annotated ORPHAcode. Among the 96 ORPHAcodes represented across the 139 patients, 18 corresponded to Subtype of disorder level. Regardless of the Orphanet classification considered, a Disorder level always has the same subtype(s) of disorder. Therefore, here again, for these 18 ORPHAcodes, we have considered their parent Disorder level, resulting in 78 unique Disorder level ORPHAcodes.

This curated set was used as the ground-truth dataset, based on two criteria: (i) diagnostic accuracy and expert validation: the expert validation was used as the confirmed ORPHAcode; (ii) cohort comprehensiveness and heterogeneity: the Solve-RD clinicians contributing these patients are affiliated with four European Reference Networks, covering major RDs clusters and ensuring that the cohort reflects the phenotypic and diagnostic diversity of the target population.

### Removing patients’ HPO redundant terms

It has been well argued that information overlap and/or imprecise clinical descriptions could lead to over-estimation of relatedness or introduce noise in ranking accuracy [21,22]. To try to avoid this issue we decided to remove parent terms if its child is also found in the annotations. To apply such a principle, we used the *minimal_set* method from the *ontologyIndex* R package, part of the OntologyX suite [23], which provides functions for ontology-aware operations. This method removes ancestor terms while retaining their descendants by traversing the HPO directed acyclic graph. For instance, when both “Microcephaly” (HP:0000252) and its parent “Abnormality of head” (HP:0000234) appear together, only the more specific child term is kept. Applying this approach across 139 patient profiles eliminated 993 redundant annotations from 2,761 total unique HPO terms.

### Measuring semantic similarity between ontology terms

Ontologies provide a formal representation of domain knowledge by organising concepts into structured hierarchies, often modelled as directed acyclic graphs in which nodes may have multiple parent and child relationships. Semantic similarity measures quantify the relatedness between ontology terms based on their positions in the graph, commonly using information content (IC), which reflects term specificity derived from annotation frequency. The IC of a term 𝑥 is defined as 𝐼𝐶(𝑥) = −𝑙𝑜𝑔(𝑝(𝑥)), where 𝑝(𝑥) is the probability of observing 𝑥 in the annotation corpus. This probability is computed as the number of occurrences of 𝑥 and its descendants in the annotations divided by the total occurrences of all terms. The root term appears in all annotations, giving it an IC of 0. More specific terms have higher IC values. When comparing two terms, IC-based approaches rely on their Most Informative Common Ancestor (MICA), defined as the shared ancestor with the highest IC.

For our study, we benchmarked six established IC-based similarity measure approaches for comparing ontology terms: Resnik [24] which uses only the IC of the MICA; Lin [25] and Jiang-Conrath (JC) from HPOSim [26], derived from Jiang-Conrath distance [27] which both incorporate the IC of individual terms and their MICA; Graph information content (GraphIC) [26] and information coefficient (SimIC) [28] which further adjust similarity measures to account for graph structure and shared IC and finally Relevance [29] which extends Lin’s measure by weighting similarity according to the probability of the MICA.

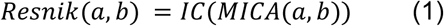

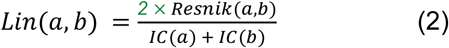

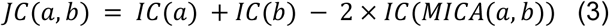

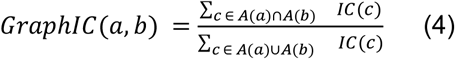

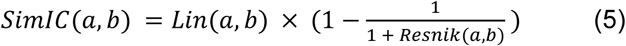

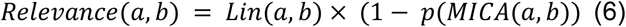

### Aggregating pairwise similarity measures

When comparing sets of terms, pairwise similarities are aggregated into a single groupwise semantic similarity metric using an aggregation function. Let 𝑋 = {𝑥_1_, 𝑥_2_, …, 𝑥_m_} denote the set of *m* HPO terms associated with a patient’s phenotype, and 𝑌 = {𝑦_1_, 𝑦_2_, …, 𝑌_n_} denote the set of *n* HPO terms associated with a candidate rare disease. Let 𝑆 ∈ ℝᵐˣⁿ represent the pairwise similarity matrix, where element Sᵢⱼ ∈ [0,1] denotes the semantic similarity between patient term xᵢ ∈ X and disease term yⱼ ∈ Y, computed using a pairwise similarity measure (e.g., Resnik, Lin, or Jiang-Conrath).

We employed four established aggregating functions recognised as the most effective state-of-the-art methods: *Best Match Average (BMA)* [11], *FunSimAvg* [29,30], *FunSimMax* [29,30] and *FunSimMaxAsym* [31]. *BMA* computes the arithmetic mean of best matches for each term across both sets. *FunSimAvg* calculates the arithmetic mean of the average best matches from each direction. *FunSimMax* selects the maximum of the two directional averages.

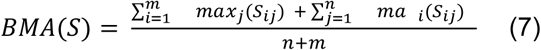

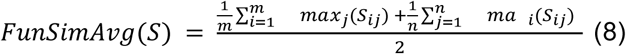

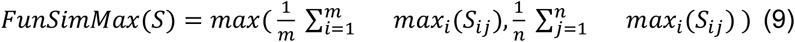

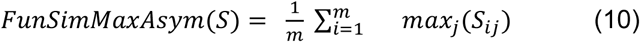

*FunSimMaxAsym* extends *FunSimMax* by aggregating semantic similarities in a single direction from patient to disease. Formally, for each patient term xᵢ ∈ X, we identify its best match among all disease terms in Y by computing maxⱼ Sᵢⱼ, then average these maximum similarities across all m patient terms. This contrasts with symmetric measures (equations 7–9) that aggregate matches in both directions, from patient to disease and from disease to patient. The asymmetric formulation prioritises complete coverage of the patient’s observed phenotype: every patient term xᵢ must find its best match within the disease’s characteristic phenotypes Y, but not all disease terms yⱼ need correspond to observed patient phenotypes. This directional choice avoids penalising diseases for possessing additional characteristic phenotypes not manifested in the patient, thereby accommodating incomplete phenotypic expression, variable disease presentation, and observational noise in clinical documentation.

### Evaluation of optimal similarity measures and aggregation methods

To identify optimal combinations of similarity measures and aggregation functions, we benchmarked all 24 combinations of the six semantic similarity measures with the four aggregation functions presented above for each of our 139 patients. For each patient, we determined the rank of their diagnosed disease among all candidate diseases and computed the mean rank across the cohort.

To average ranks, the harmonic mean emphasises smaller ranks, making it suitable for averaging ranks where lower ranks matter more than higher ones:

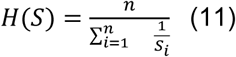

Where 𝑛 is the number of elements in the set S.

To visualise global ranking performance, we calculated and plotted cumulative distribution functions (CDFs). The CDF quantifies the probability P(X ≤ r) that the diagnosed disease ranks at position r or better, providing a global view of ranking effectiveness across the cohort.

### HPO term frequency weighting and excluded terms

We hypothesised that highly prevalent phenotypes within a disease should weigh more heavily in similarity computations than rare phenotypes. To evaluate this, as applied in the Dream Rare-X project [32], we mapped the frequency categories, excepting the “excluded” one, to numerical weights using monotonic vectors of length five, where each position corresponds to one category in descending frequency order. Hence, we generated 31 weighting schemes: (5,4,3,2,1), (4,4,3,2,1), (4,3,3,2,1), …, (2,1,0,0,0), (1,1,0,0,0), (1,0,0,0,0), testing all patterns where weights decrease or remain constant from “obligate” to “very rare” categories. We modified the aggregation function as follows:

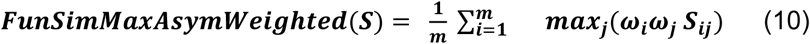

where 𝜔_i_ and 𝜔_j_ represent the frequency-derived weight for the terms x_i_ and x_j_.

Clinical signs annotated as “excluded”, indicating phenotypes incompatible with a diagnosis, triggered automatic removal of the corresponding disease from ORPHAcode candidates list.

### Random-walk propagation on disease similarity network

We constructed a network in which nodes represent RD and edges their phenotypic similarity. For each pair of RD, we computed a single groupwise similarity measure based on HPO annotations using the symmetric aggregation function FunSimMax.

To prioritise candidate RD for a given patient, we extended this network by adding a patient node connected to each RD. The weight of each patient-RD edge was computed using the asymmetric function FunSimMaxAsym.

We then applied a Random Walk with Restart propagation on this network [33,34]. At each step the walker either follows an outgoing edge according to the transition probabilities derived from edge weights or restarts to the patient node with a given probability. The walk converges to a stationary distribution, which we interpret as a similarity score, with higher values indicating greater phenotypic relatedness to the patient.

The damping factor α controls the balance between exploration and restart. Higher values of α enable longer walks and global propagation, while lower values increase restart frequency and emphasize local neighbourhoods.

By constraining the walk through an appropriately low damping factor α, we maintain locality around the patient node. This constraint limits the disproportionate influence of highly connected hubs, which can otherwise introduce bias into the ranking process. This local exploration prioritises clinically relevant RD candidates by integrating: (i) direct similarity between patient and RD and (ii) indirect phenotypic relationships mediated through the RD-RD similarity network structure.

### Technical implementation

Our framework is implemented in the Python Programming Language version 3.12 and takes advantage of Python’s extensive collection of data science libraries. In particular, pandas 2.2 is used for data manipulation and NetworkX v3.4 for network-related tasks. Ontology queries and HPO term inspection were performed with the hpo3 library (v1.4), which includes the March 2025 HPO release, and ontologyX 2024-02-20 R package for the subsuming HPO section. The workflow is executed in a Linux environment (Ubuntu 22.04) and organised into a reproducible workflow using Snakemake 9.3. To ensure full reproducibility, the repository provides a Conda 25.3 environment. The Snakemake workflow automates the entire pipeline, including semantic similarity computation, network construction, random-walk execution, and ranking evaluation results. All source code and documentation required to reproduce the analyses presented in this study are available at https://github.com/Orphanet/OrphaScape_SimWalk

### Data availability

The Solve-RD subset analysed during the current study is based on datasets available as phenopackets at the EGA (EGAD00001009767, EGAD00001009768, EGAD00001009769 and EGAD00001009770 under Solve-RD study EGAS00001003851).

## Results

### A phenotype-based pipeline for rare disease diagnosis guidance

Following up the Solve-RD project [7], we developed a phenotype-based pipeline to guide rare disease diagnosis (Fig. 1). The pipeline takes three inputs: the HPO ontology, the Orphadata *’Phenotypes Associated with Rare Disorders’* dataset and a patient’s HPO-encoded phenopacket (Fig. 1a). It computes phenotypic similarity between the patient and ORPHAcodes (Fig. 1d), then constructs a weighted network where nodes represent diseases and the patient, with edge weights reflecting phenotypic similarity (Fig. 1e). Random walk with restart (RWR) propagates information from the patient node through this network, yielding a ranked list of candidate ORPHAcodes (Fig. 1g). Users can export the output network to graph database systems for visualisation and analysis. This enables exploration of the patient’s local neighbourhood, including disease-associated genes, to guide subsequent variant investigation (Fig. 1h).

**Fig. 1.**
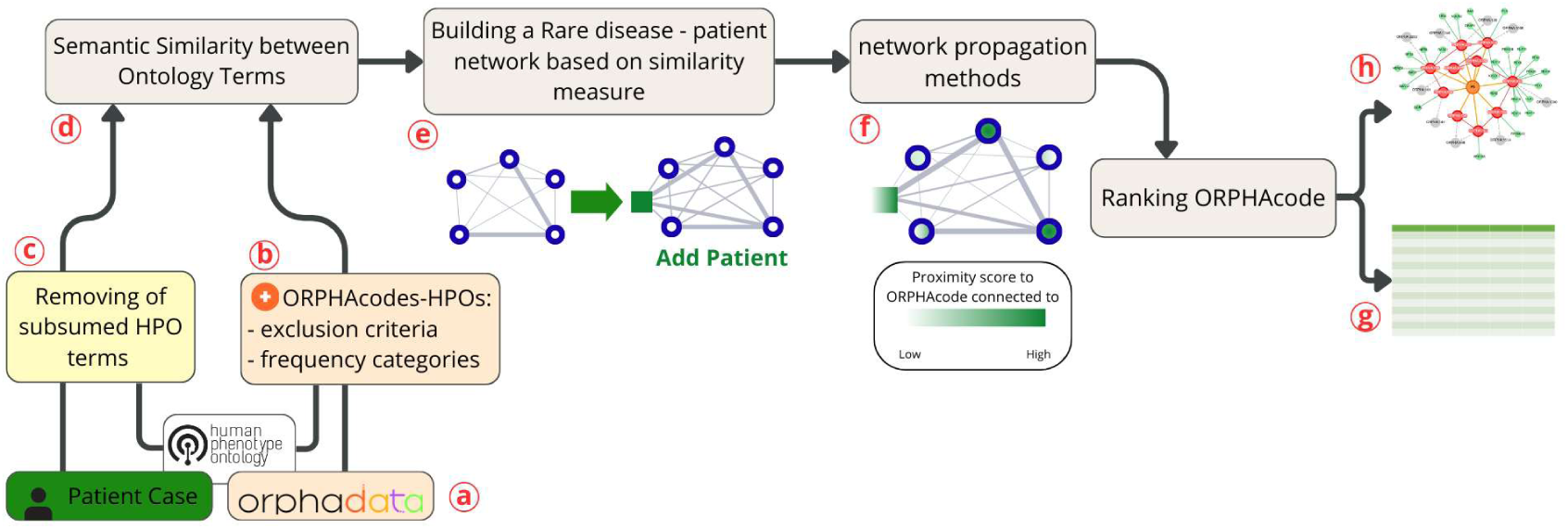
Overview of the phenotype-based pipeline.

Before computing semantic similarity, we applied two pre-processing steps (Fig. 1b-c): firstly, we incorporated estimated frequency categories for each HPO term associated with a disease from Orphanet data, and secondly we removed subsumed HPO terms from patient profiles when more specific descendant terms were present.

To determine the optimal semantic similarity approach (Fig. 1d), we benchmarked all 24 combinations of six IC-based similarity measures with four groupwise aggregation methods, all described in the Methods section. Each combination produces a single similarity score between the patient and each ORPHAcode.

### Resnik + FunSimMaxAsym outperforms other groupwise semantic similarity measures

We evaluated 24 configurations combining six pairwise similarity measures (Resnik, Lin, Jiang-Conrath, Relevance, Information Coefficient, GraphIC) with four aggregation functions (BMA, FunSimAvg, FunSimMax, FunSimMaxAsym) across the 139-patient cohort. Figure 2 presents cumulative distribution functions for the seven highest-performing configurations alongside the RSD baseline. Resnik + FunSimMaxAsym (red) consistently ranked confirmed diagnoses lowest, placing 38% of correct ORPHAcodes within the top 10 positions compared with 29% for RSD (blue). This configuration achieved a harmonic mean rank of 5.45 versus 7.97 for RSD (Table 1). Complete results for all 24 configurations appear in Supplementary Table S1.

**Fig. 2.**
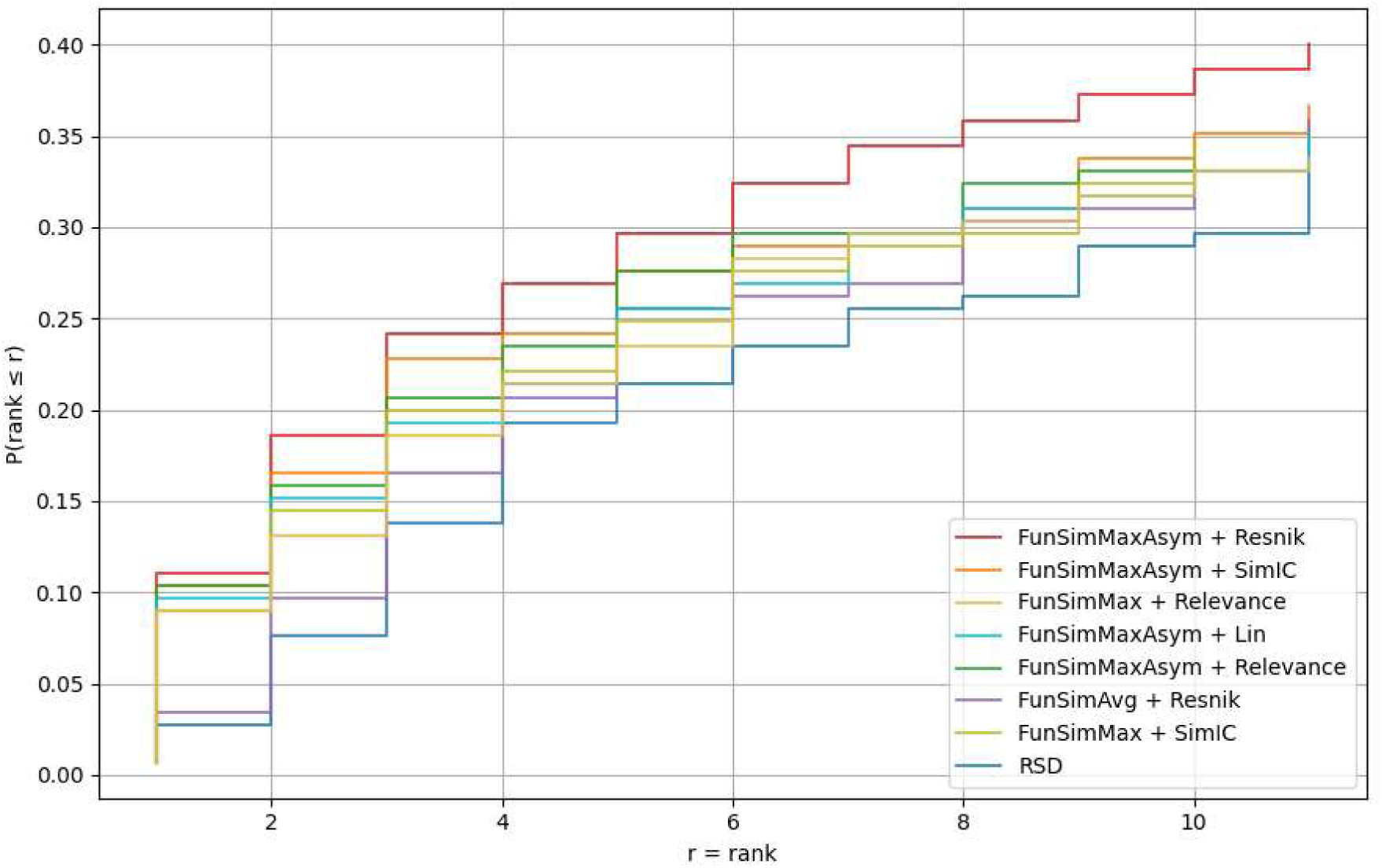
Performance comparison of semantic similarity measures and aggregation methods for ranking rare disease diagnoses. Cumulative distribution functions for the seven highest-performing configurations (out of 24) and the RSD baseline (blue) across 139 patients. The x-axis represents the rank position of the correct ORPHAcode; the y-axis shows the cumulative proportion of patients whose correct diagnosis appears at or above that rank. CDFs positioned towards the upper-left indicate superior performance.

**Table 1.** Harmonic mean ranks of correct diagnoses across semantic similarity configurations (7 best + RSD). Performance of 7 best combinations of pairwise semantic similarity measures and aggregation methods, plus the RSD baseline, evaluated across 139 patients with 78 distinct confirmed ORPHAcodes. Lower harmonic mean ranks indicate superior diagnostic ranking performance. Complete results for all 24 configurations appear in Supplementary Table S2.

| Pairwise semantic similarity measure | Aggregation method | Harmonic mean rank |
| --- | --- | --- |
| Resnik | FunSimMaxAsym | 5.45 |
| Simlc | FunSimMaxAsym | 5.85 |
| Relevance | FunSimMaxAsym | 5.96 |
| Lin | FunSimMaxAsym | 6.26 |
| Simlc | FunSimMax | 6.37 |
| Relevance | FunSimMax | 6.52 |
| Resnik | FunSimAvg | 7.81 |
| RSD |  | 7.97 |

### Effect of subsumed HPO terms removal on ranking

Figure 3 compares ranking performance with (green) and without (red) subsumed terms using Resnik + FunSimMaxAsym. Filtering subsumed terms retrieves the confirmed ORPHAcode within the top ten positions for 38% of patients, compared with 35% before filtering and 25% for the RSD baseline (blue).

**Figure 3.**
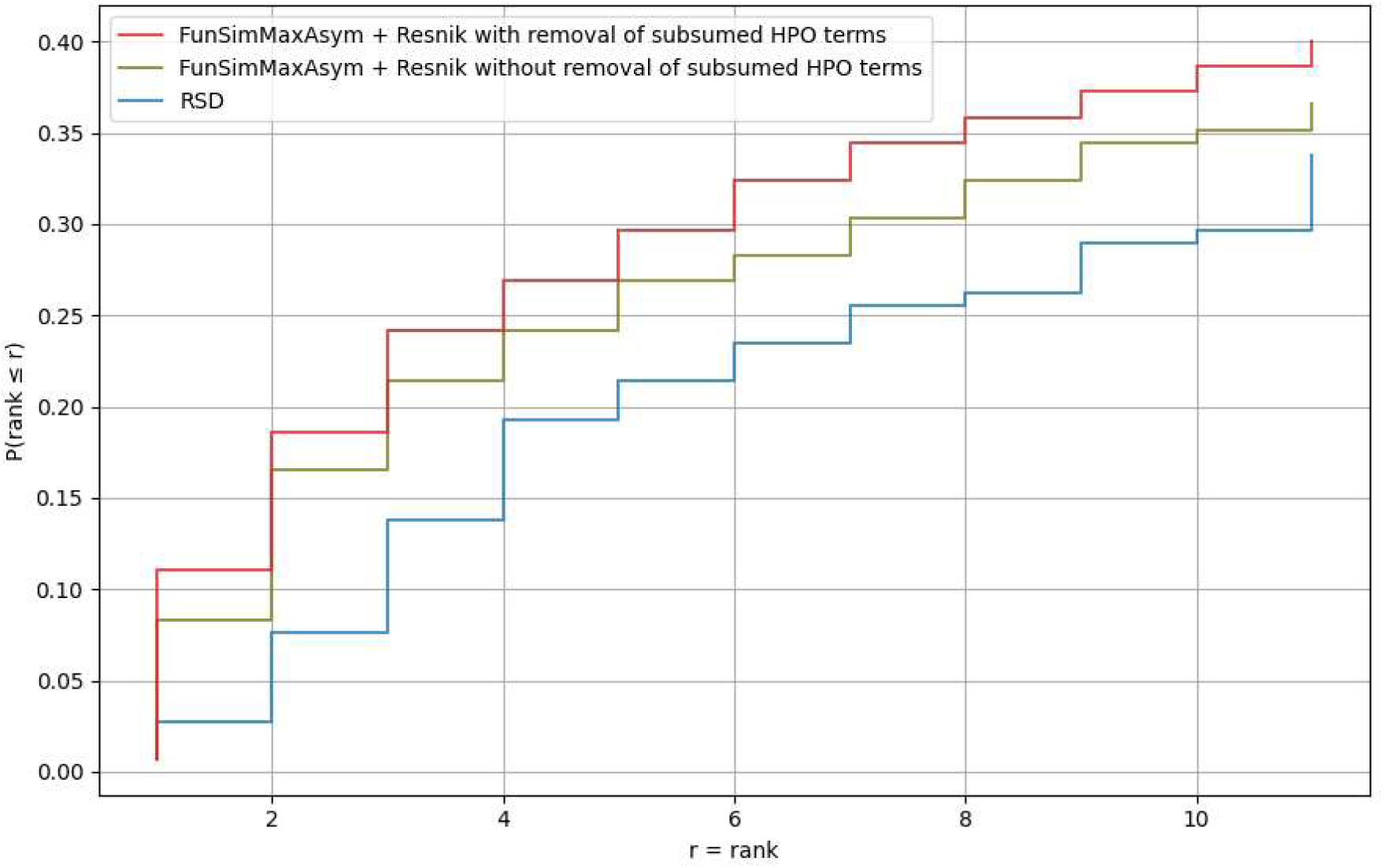
Diagnostic ranking performance with and without subsumed term filtering. Comparison of Resnik + FunSimMaxAsym using unfiltered (green) versus filtered (red) patient profiles, and RSD baseline (blue). The cumulative distribution function shows the proportion of patients whose confirmed diagnosis ranked at position r or better.

Removing subsumed HPO terms improves ranking performance. As shown in Table 2, the harmonic mean rank of the confirmed ORPHAcode decreases from 5.45 to 4.73 after filtering (lower is better).

**Table 2.** Harmonic mean rank of confirmed ORPHAcodes before and after filtering subsumed HPO terms. Lower values indicate better performance.

| Method | Harmonic mean rank |
| --- | --- |
| Removal | 4.73 |
| No removal | 5.45 |
| RSD | 7.97 |

This filtering strategy is applied exclusively to patient profiles. In contrast, HPO annotated ORPHAcodes are not filtered because they represent population-level descriptions with frequency estimates assigned to terms at multiple granularity levels. Removing subsumed terms would eliminate this frequency information and reduce the specificity of disease annotations.

### Effect of frequency-based weighting on ranking performance

We developed and evaluated a weighted variant of *FunSimMaxAsym* where HPO terms are weighted according to their annotated frequency in the five categories involved: obligate, very frequent, frequent, occasional, very rare (see Materials and Methods).

Figure 4 and Table 3 suggest that down-weighting very rare clinical signs improves ranking performance. Given our limited sample size, these represent preliminary trends. The weighting scheme (2,2,2,2,1), which reduces the contribution of very rare signs (to half the weight of other categories), performed best, followed by (1,1,1,1,0), which excludes very rare terms entirely.

**Figure 4.**
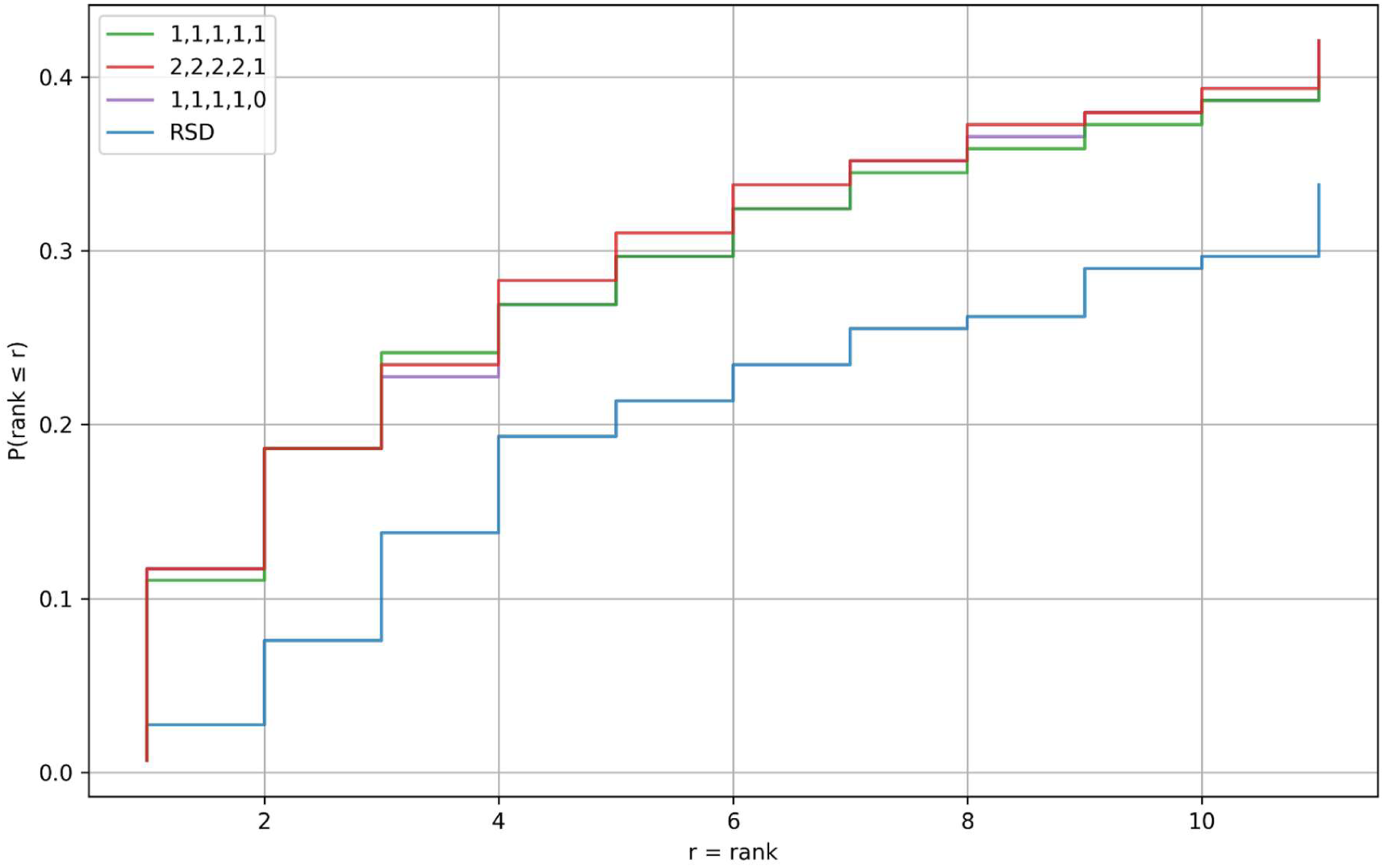
Impact of frequency-based weighting on diagnostic ranking performance. Cumulative distribution functions comparing four approaches: three frequency-weighting schemes for *FunSimMaxAsym*: (2,2,2,2,1) (red), (1,1,1,1,0) (purple), unweighted control (1,1,1,1,1) (green), and the RSD baseline method (blue). The weighting vectors correspond to the five Orphanet frequency categories in descending order: obligate, very frequent, frequent, occasional, very rare.

**Table 3.** Harmonic mean rank of correct diagnoses under frequency-weighting schemes. Performance comparison of four approaches: three frequency-weighting variants of FunSimMaxAsym: (2,2,2,2,1), (1,1,1,1,0), and unweighted control (1,1,1,1,1), and the RSD baseline method. Weighting vectors correspond to the five Orphanet frequency categories in descending order: obligate, very frequent, frequent, occasional, very rare. Lower harmonic mean ranks indicate superior diagnostic ranking performance. Complete results for all 31 tested weighting schemes are provided in the Supplementary Table S3.

| Weighting | Harmonic mean rank |
| --- | --- |
| 2,2,2,2,1 | 4.64 |
| 1,1,1,1,0 | 4.69 |
| 1,1,1,1,1 | 4.73 |
| RSD | 7.97 |

### Effect of network propagation on clinical consistency of top-ranked candidates

Rather than focusing only on changes in the average rank of the confirmed diagnosis before and after applying RWR (Figure S1), we evaluated whether network propagation yields clinically more coherent candidate rankings according to the Orphanet nomenclature. Specifically, we assessed RWR’s ability to improve clinical consistency amongst the candidates. We conducted two complementary analyses to address this question.

Both analyses evaluated clinical consistency by counting the number of GDs shared between each candidate ORPHAcode and the confirmed ORPHAcode, but differed in their scope of hierarchy traversal.

In the first analysis, we considered, for each disease, only its immediate parent GD (i.e. the most specific GD) across all Orphanet classifications in which it appears. We computed the harmonic mean rank of candidate ORPHAcodes sharing at least one of these immediate parent GDs with the confirmed diagnosis. This horizontal approach in Orphanet classifications captures the diversity of clinical contexts in which a disease may be classified, without the constraint of a single preferential hierarchy, whilst limiting depth to the most specific classification level.

The damping factor α substantially influenced performance, with optimal results at α ∈ [0.25, 0.5]. At α = 0.3, diseases sharing the relevant GDs achieved a harmonic mean rank of 14.13, compared with 16.7 before propagation (α = 0) and 25.57 for RSD (Table 4).

**Table 4:** Harmonic mean rank of ORPHAcodes sharing at least one lowest-level GD with the confirmed ORPHAcode across varying damping factor α values.

| Damping factor value | Harmonic mean rank of ORPHAcodes with shared GDs |
| --- | --- |
| *RSD* | 25.57 |
| 0 (no random walk) | 16.70 |
| 0.05 | 17.50 |
| 0.1 | 17.50 |
| 0.15 | 17.63 |
| 0.2 | 17.74 |
| 0.25 | 14.14 |
| **0.3** | **14.13** |
| 0.35 | 14.23 |
| 0.4 | 14.32 |
| 0.45 | 14.51 |
| 0.5 | 14.53 |
| 0.6 | 17.27 |
| 0.7 | 20.19 |
| 0.8 | 30.84 |
| 0.9 | 120.09 |
| 1.0 | 722.76 |

In the second analysis, we restricted our approach to the preferential parent hierarchy of each disease, considering all GDs within this single classification branch. This vertical approach captures clinical consistency along the most relevant classification path for each disease, avoiding noise from distantly related alternative classifications whilst maintaining rich clinical information across multiple hierarchical levels within that branch.

RWR increased the proportion of patients with at least one shared GD between top 10 candidates and confirmed diagnosis from 14% (α = 0) to 30% (α = 0.3). Patients with multiple shared GDs increased from 0.7% to 13%. Table 5 presents detailed GDs overlap counts for 10 representative patients with highest baseline overlap, demonstrating that 9 out of 10 showed increased GDs sharing after RWR. Across the full cohort, 16% of patients showed improved GD overlap following propagation.

**Table 5.** Top ten patients with the highest number of distinct shared GDs between their top ten ranked ORPHAcode candidates and the confirmed ORPHAcode, restricted to the preferential parent classification. Patients are ranked based on the number of GD prior to applying the RWR.

| Patient and confirmed<br>ORPHAcode | Shared GDs within top-10<br>candidates (pre-RWR) | Shared GDs within top-10<br>candidates (post-RWR) |
| --- | --- | --- |
| P3294 (ORPHA:199) | 8 | 19 |
| P6438 (ORPHA:90652) | 8 | 6 |
| P6774 (ORPHA:1465) | 7 | 12 |
| P6777 (ORPHA:1465) | 7 | 9 |
| P3237 (ORPHA:442835) | 6 | 7 |
| P3837 (ORPHA:2332) | 5 | 7 |
| P5345 (ORPHA:2020) | 4 | 5 |
| P8285 (ORPHA:2152) | 3 | 6 |
| P9601 (ORPHA:36386) | 2 | 4 |
| P8535 (ORPHA:352577) | 2 | 3 |

Both analyses demonstrate that RWR enhances clinical consistency: the ORPHAcodes sharing lowest-level DG with confirmed diagnoses rank higher within top 10 predictions after network propagation and among these, the ORPHAcodes sharing the highest number of GD restricted to the preferential parent come from after applying the RWR, providing clinically interpretable candidate clusters that facilitate differential diagnosis.

### Effect of network propagation on diagnostic performance

We evaluated damping factor (α) values to assess the trade-off between individual diagnosis ranking and clinical consistency of top predictions. Increasing α progressively decreased ranking performance for confirmed ORPHAcodes. As previously mentioned, at α=0 (no network propagation), the method achieved a harmonic mean rank of 4.64. Applying RWR with α=0.3 yielded a harmonic mean rank of 5.55, while α values above 0.7 performed worse than the RSD baseline (7.97).

Despite lower individual ranking performance, α=0.3 maximized clinical consistency by prioritizing ORPHAcodes that share GDs with the confirmed diagnosis within the top 10 predictions, where shared GDs were restricted to the preferential parent hierarchy (see clinical consistency analysis). This configuration balances two objectives: maintaining competitive ranking (5.55 vs 4.64 without RWR) while enriching top predictions with clinically related diseases that aid differential diagnosis. For applications prioritising strict ranking accuracy, α=0 remains optimal; for clinical decision support requiring interpretable disease candidates, α=0.3 provides superior utility. Complete results for all α values appear in Supplementary Table S1.

## Discussion

This study demonstrates that combining phenotype-based semantic similarity with network propagation effectively ranks rare disease diagnostic hypotheses and promotes clinical consistency among top candidates according to Orphanet nomenclature. Three methodological refinements contributed to performance: first, aggregating pairwise Resnik similarities using FunSimMaxAsym provided robust patient-disease comparison; second, removing subsumed HPO terms reduced ontological redundancy, yielding more discriminative phenotype representations; third, down-weighting very rare clinical features refined ranking by emphasising characteristic disease features.

### Network propagation enhances clinical consistency through group of disorders alignment

Random walk with restart on the ORPHAcode similarity network leverages both direct phenotypic matches and indirect relationships through shared network topology. This approach addresses a fundamental limitation of direct similarity measures: annotation density bias. Orphanet’s phenotypic annotations are manually curated by clinical experts and maintain high individual quality. However, annotation completeness varies across diseases due to three factors: (i) the period when annotation was performed, (ii) available medical knowledge and case reports at that time, and (iii) the HPO release version used. Despite regular updates, diseases with sparse annotations may rank poorly even when clinically relevant, while extensively annotated diseases tend to rank highly regardless of appropriateness (31).

RWR mitigates this bias by exploring the local neighbourhood around the patient node, identifying indirectly related diseases through network connectivity rather than direct annotation overlap. By leveraging Orphanet’s classification hierarchy, RWR compensates for incomplete annotations and captures phenotypically homogeneous disease clusters. Within the top 10 predictions, RWR consistently prioritised diseases sharing many lowest-level GDs with confirmed diagnoses or exhibiting the same GDs when constrained to preferential parent classifications (Table 4). This clinical consistency proves particularly valuable: although individual confirmed diagnoses occasionally ranked lower, their GD neighbours appeared prominently, maintaining diagnostic utility. The dual benefits of RWR, exploiting indirect node similarities and reinforcing topologically coherent candidates, address challenges from phenotypic heterogeneity and incomplete patient presentations that challenge purely similarity-based approaches.

These findings support clinical decision support systems that present disease clusters rather than isolated predictions, improving interpretability when clinicians evaluate differential diagnoses for rare diseases. Ongoing efforts at Orphanet focus on improving annotation depth and consistency, with emerging Retrieval-Augmented Generation techniques showing promise for enhancing annotation accuracy and workflow efficiency.

### Frequency weighting and exclusion criteria enhance accuracy despite coverage limitations

Incorporating Orphanet frequency annotations improved ranking performance (Table 3, Fig. 4). The optimal weighting scheme (2,2,2,2,1) down-weighted very rare phenotypes while preserving contributions from obligate through occasional features, thereby emphasising characteristic disease patterns over nonspecific findings.

Orphanet also flags certain phenotypes as “excluded,” indicating clinical signs whose presence precludes a diagnosis. Our pipeline integrates these exclusion criteria by automatically removing incompatible diseases from candidate lists. However, annotation coverage remains limited: fewer than 400 of approximately 4,300 annotated ORPHAcodes (8.05%) contain exclusion flags, and excluded phenotypes represent only 0.62% of all HPO annotations (see proportions in Supplementary Information S4). Although Orphanet continues expanding these annotations, current sparsity constrains their diagnostic impact. However, this small number of exclusion annotations does not stand for a weakness in the annotation process but arises because relatively few RDs require the exclusion of a phenotype to establish a valid diagnosis. This is due to the phenotypic variability of RDs, and it is very difficult to assert that a clinical sign never occurs in a specific RD, except in a relatively small number of cases.

### Structural challenges in ontology-based similarity computation

Ontology-based similarity measures face inherent challenges when applied to HPO. First, HPO’s multiple inheritance structure, where terms belong to several branches, introduces path-length variability that influences similarity scores inconsistently across the ontology [35]. Second, sibling terms sharing a parent node receive high similarity scores despite potentially opposing clinical meanings; for example, Hypertonia (HP:0001276) and Hypotonia (HP:0001252) both descend from Abnormal muscle tone (HP:0003808) yet represent contradictory phenotypes [36]. Such structural ambiguities can produce misleading similarity scores that do not reflect clinical reality.

Our approach partially mitigates these limitations: removing subsumed terms reduces path-length effects; FunSimMaxAsym’s patient-centred directionality focuses on observed phenotypes rather than complete disease profiles; network propagation supplements phenotypic similarity with disease group topology. However, fundamental ontological ambiguities remain unresolved. The requirement for patients to present at least five clinical signs [22,37] addresses data sparsity rather than structural issues, though richer phenotypic profiles do improve discriminative power.

These limitations underscore that purely phenotype-based approaches cannot fully capture diagnostic complexity. Integrating additional information sources (genetic data, temporal progression, or biochemical markers) represents a necessary direction for robust rare disease decision support systems.

### Limitations

Phenotype-based similarity measures depend critically on the completeness and specificity of clinical annotations especially for patients. Our ground-truth cohort from Solve-RD exhibits several feature biases that may affect generalisability. First, the historical annotation protocol restricted clinicians to a predefined HPO subset, limiting representation of phenotypic heterogeneity. Second, patient recruitment through specific European Reference Networks may have introduced domain-specific HPO usage patterns that inadequately capture the multisystemic nature of many rare diseases. Third, individual clinician expertise likely introduced variation in term selection and granularity. These biases may contribute to suboptimal classification of confirmed diagnoses in cases where patient phenotyping differs from Orphanet annotations, which are made on the basis that in scientific literature a population of patients presents a certain clinical sign(s) with a specified occurrence.

The cohort size (139 patients representing 78 distinct ORPHAcodes) further limits statistical power for detecting performance differences between methods, particularly for rare weighting schemes and damping factor configurations. External validation on independent cohorts with different annotation workflows is required to assess generalisability. Future annotation strategies should encourage cross-specialty, multi-domain phenotyping to better reflect the complexity of rare disease presentations and improve diagnostic tool performance.

Beyond addressing these current limitations, future work will integrate genetic data, including disease-associated genes and variant information, to guide candidate prioritisation within patients’ network neighbourhoods, enabling combined phenotypic-genomic diagnostic workflows that address the inherent limitations of purely phenotype-based approaches identified in this study.

## Supporting Information

**S1 Table.**
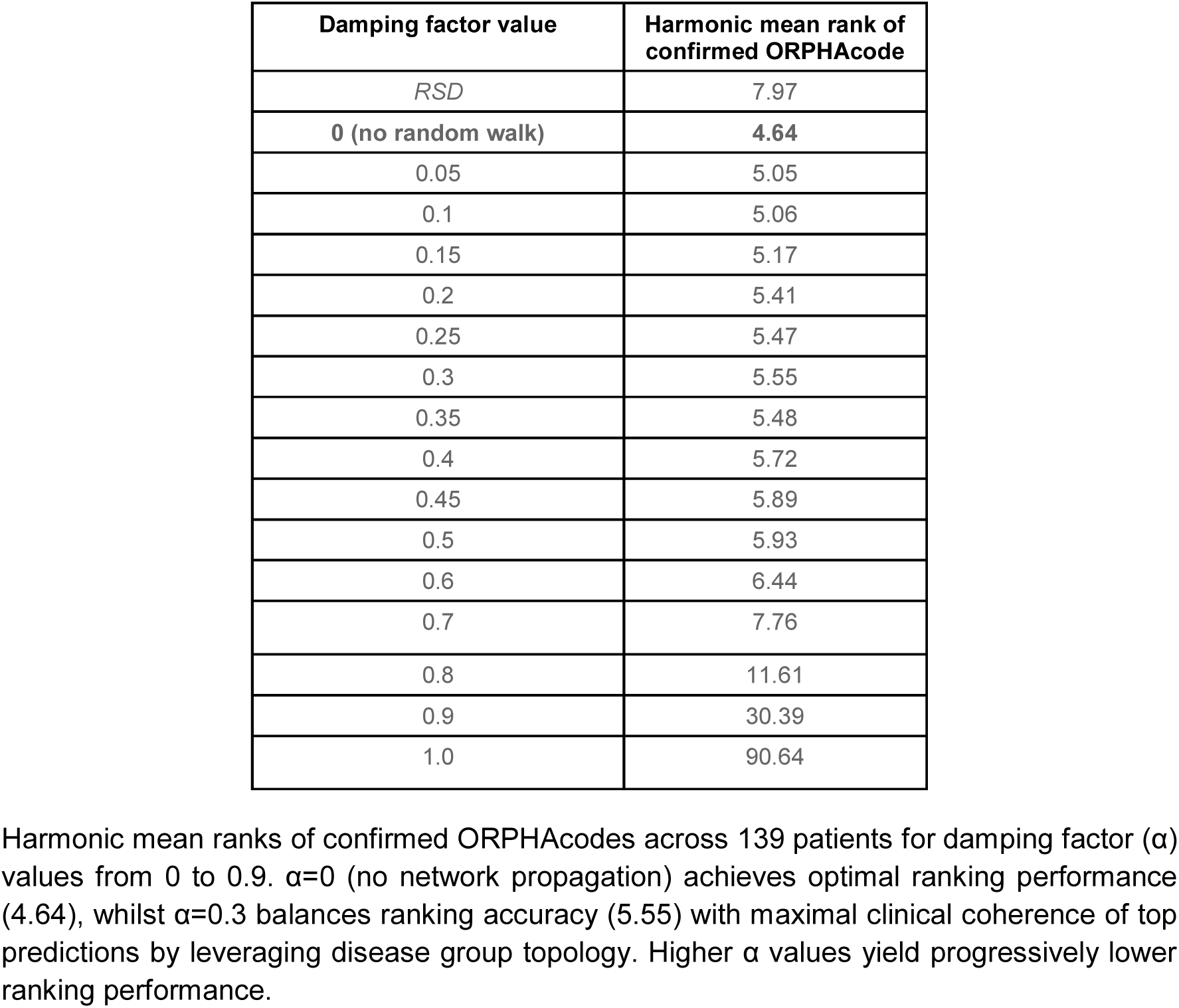
Effect of damping factor on ranking performance.

**S2 Table.** Harmonic mean ranks of correct diagnoses across semantic similarity configurations (7 best + RSD).

| Pairwise semantic similarity measure | Aggregation method | Average rank |
| --- | --- | --- |
| **Resnik** | **FunSimMaxAsym** | **5.45** |
| Simlc | FunSimMaxAsym | 5.85 |
| Relevance | FunSimMaxAsym | 5.96 |
| Lin | FunSimMaxAsym | 6.26 |
| Simlc | FunSimMax | 6.37 |
| Resnik | FunSimMax | 6.43 |
| Relevance | FunSimMax | 6.52 |
| Relevance | FunSimAvg | 6.65 |
| Simlc | FunSimAvg | 6.87 |
| Lin | FunSimMax | 7.00 |
| GraphIC | FunSimMaxAsym | 7.17 |
| Lin | FunSimAvg | 7.35 |
| JC | FunSimMaxAsym | 7.54 |
| Resnik | FunSimAvg | 7.81 |
| RSD |  | 7.97 |
| GraphIC | FunSimMax | 8.33 |
| Lin | FunSimMax | 8.37 |
| GraphIC | FunSimAvg | 8.78 |
| JC | FunSimAvg | 8.95 |
| JC | BMA | 15.63 |
| Simlc | BMA | 15.72 |
| Lin | BMA | 15.91 |
| Relevance | BMA | 16.10 |
| Resnik | BMA | 16.44 |
| GraphIC | BMA | 16.51 |

Performance of 24 combinations of pairwise semantic similarity measures (Resnik, Lin, Jiang-Conrath, Relevance, InformationCoefficient, GraphIC) and aggregation methods (BMA, FunSimAvg, FunSimMax, FunSimMaxAsym), plus the RSD baseline, evaluated across 139 patients with 78 distinct confirmed ORPHAcodes. Lower harmonic mean ranks indicate superior diagnostic ranking performance.

**S3 Table.** Harmonic mean rank of correct diagnoses under frequency-weighting schemes (full data)

| Weighting | Average rank |
| --- | --- |
| **2,2,2,2,1** | **4.64** |
| 1,1,1,1,0 | 4.69 |
| 1,1,1,1,1 | 4.73 |
| 3,2,2,2,1 | 4.78 |
| 3,3,3,2,1 | 5.47 |
| 4,3,3,2,1 | 5.52 |
| 2,1,1,1,0 | 5.61 |
| 2,2,2,1,0 | 5.62 |
| 2,2,2,1,1 | 5.64 |
| 2,1,1,1,1 | 5.66 |
| 1,1,1,0,0 | 5.78 |
| 3,2,2,1,0 | 5.80 |
| 3,2,2,1,1 | 5.81 |
| 3,3,2,2,1 | 6.23 |
| 4,3,2,2,1 | 6.28 |
| 4,4,3,2,1 | 6.75 |
| 5,4,3,2,1 | 6.84 |
| 2,1,1,0,0 | 6.99 |
| 3,3,2,1,1 | 7.07 |
| 4,3,2,1,1 | 7.10 |
| 3,3,2,1,0 | 7.23 |
| 4,3,2,1,0 | 7.24 |
| 2,2,1,1,1 | 7.37 |
| 3,2,1,1,1 | 7.60 |
| 2,2,1,1,0 | 7.96 |
| RSD | 7.97 |
| 3,2,1,1,0 | 8.25 |
| 2,2,1,0,0 | 8.33 |
| 3,2,1,0,0 | 8.77 |
| 1,1,0,0,0 | 11.59 |
| 2,1,0,0,0 | 18.32 |
| 1,0,0,0,0 | 33.90 |

**S4.a :** Proportion of HPO term frequency categories across all 115767 ORPHAcode’s HPO term annotations.

| HPO term frequency | Number HPO annotations | Proportion (%) |
| --- | --- | --- |
| Obligate (100%) | 624 | 0.54 |
| Very frequent (99-80%) | 25698 | 22.20 |
| Frequent (79-30%) | 39412 | 34.04 |
| Occasional (29-5%) | 42764 | 36.94 |
| Very rare (<4-1%) | 6550 | 5.66 |
| Excluded (0%) | 719 | 0.62 |

**S4.b:** Proportion of HPO term frequencies across all 4319 annotated ORPHAcodes from the “Phenotypes Associated with Rare Disorders” May 2025 release.

| HPO term frequency | Number of ORPHAcodes | Proportion (%) |
| --- | --- | --- |
| Obligate (100%) | 356 | 8.24 |
| Very frequent (99-80%) | 3651 | 84.53 |
| Frequent (79-30%) | 3831 | 88.70 |
| Occasional (29-5%) | 3426 | 79.32 |
| Very rare (<4-1%) | 1093 | 25.31 |
| Excluded (0%) | 348 | 8.06 |

## Acknowledgements

The Solve-RD project has received funding from the European Union’s Horizon 2020 research and innovation programme under grant agreement No 779257.

The authors gratefully acknowledge Anaïs Baudot, who reviewed this manuscript and provided insightful feedback, and Jonas Marcello for his valuable technical guidance and advice regarding the hpo3 library.

